# GLP-1 receptor agonist indications and prescriptions among adults who do and do not smoke

**DOI:** 10.1101/2025.08.12.25331941

**Authors:** Eleanor L.S. Leavens, Michael J. Arnold, Nicole L. Nollen, Lisa Sanderson Cox, Kristy A. Brown, Edward F. Ellerbeck

**Author notes:** **Corresponding author:** Eleanor L. S. Leavens, PhD, Assistant Professor, Department of Population Health, University of Kansas Comprehensive Cancer Center, Kansas City, KS 66160.

## Abstract

**Background:** Smoking and obesity are leading causes of preventable morbidity and mortality, often co-occurring and synergistically increasing risks for cardiovascular disease and diabetes. Glucagon-like peptide-1 receptor agonists (GLP-1 RAs) are effective for managing type 2 diabetes, obesity, and cardiovascular disease, and emerging evidence suggests potential benefit for smoking cessation. However, the prevalence of GLP-1 RA indications and prescriptions among adults who smoke is unknown.

**Objective:** To compare the frequency of GLP-1 RA indications and prescriptions among adults who do and do not smoke within a large Midwest health system.

**Methods:** Electronic health record data identified adults (≥18 years) with outpatient primary care visits from September 2023–2024. Patients were categorized as current smokers or non-smokers. We assessed indications for GLP-1 RAs—type 2 diabetes, weight management (BMI ≥30 or 27– 29.9 with comorbidity), and secondary prevention of cardiovascular disease and examined prescription rates among those with at least one indication.

**Results:** Of 59,981 patients, 4,843 (8.1%) were current smokers. Compared to non-smokers, adults who smoke were more likely to have an indication for GLP-1 RAs (66.1% vs. 62.6%; *p*<0.001) but less likely to receive a prescription (20.4% vs. 23.2%; *p*<0.001). This pattern was consistent across all indications.

**Conclusion:** Adults who smoke more frequently meet clinical criteria for GLP-1 RAs yet are less likely to be prescribed these therapies. Enhancing access to GLP-1 RAs for individuals who smoke could address critical metabolic and smoking-related health challenges, particularly given their potential role in supporting smoking cessation.

## BACKGROUND

Smoking and obesity are the two leading causes of preventable death and disease in the United States. The two conditions, which often co-occur, are synergistically detrimental to health.^1,2^ Individuals who smoke and have obesity are at greater risk for cardiovascular disease and diabetes, while also experiencing reduced quality of life and greater healthcare costs compared to individuals with either condition alone.^3,4^ Glucagon-like peptide-1 receptor agonists (GLP-1 RAs) are widely used for the treatment of type 2 diabetes, obesity, and cardiovascular disease, but research on their use for comorbid smoking is in its infancy. A 6-week pilot study of exenatide added to nicotine patch^5^ in adults with smoking and prediabetes and/or overweight reported a 46% abstinence rate - nearly double that of currently approved pharmacotherapy^5^ — along with a 5.6 pound reduction in post-cessation body weight. GLP-1 RAs are the most promising advance for individuals with both tobacco use and obesity in over two decades, but the relative prevalence of indications for and uptake of GLP-1 RAs is unknown.

## OBJECTIVE

This analysis of adults within a large Midwest health system compares the frequency of indications for GLP-1 RAs and GLP-1 RA prescriptions among adults who do and do not smoke.

## METHODS

Data from a large, Midwest health system’s electronic health record (EHR) was used to identify adult patients ≥ 18 years of age receiving care in an outpatient primary care clinic between September 2023 to September 2024. From the overall sample, we identified the proportion identified in the EHR as being a current smoker versus non-current smoker. Within those two groups, we then identified patients with a current GLP-1 RA indication including for 1) the management of diabetes mellitus, 2) weight management (BMI ≥ 30.0 or BMI 27.0-29.9 with a weight-related comorbidity [i.e., hypertension, hyperlipidemia/hypercholesterolemia, obstructive sleep apnea, diabetes mellitus]), and 3) secondary prevention of cardiovascular disease (age 45+ with BMI of 27+ and history of myocardial infarction/stroke). Of those with at least one indication, we determined the proportion prescribed a GLP-1 RA including lixisenatide, exenatide, dulaglutide, liraglutide, semaglutide, or tirzepatide.

## RESULTS

We identified 59,981 adult patients in primary care of whom 4,843 (8.1%) were current smokers. Adults who smoke were more likely to have an indication for a GLP-1 RA compared to non-smokers (66.1% vs 62.6%; p < 0.001) but were less likely than non-smokers to receive a GLP-1 RA (20.4% vs 23.2%; p < 0.001; Table 1). These findings were consistent across all indications for GLP-1 RAs, including diabetes, weight management, and cardiovascular disease prevention.

**Table 1.** Indications for and utilization of GLP-1 RA^a^ among adults who do and do not smoke.

|  | Indication for GLP-1 RA<br>(n (%)) |  |  | Prescribed GLP-1 RA<sup>b</sup><br>(n (%)) |  |  |
| --- | --- | --- | --- | --- | --- | --- |
|  | Non-smokers<br>(n=55,138) | Current smokers<br>(n=4,843) | P value | Non-smokers<br>(n=34,496) | Current smokers<br>(n=3,199) | P value |
| **GLP-1 RA<sup>c</sup> INDICATIONS** |  |  |  |  |  |  |
| **Diabetes** | 9,415 (17.1) | 1,019 (21.0) | <0.0001 | 3,329 (35.4) | 326 (32.0) | 0.02 |
| **Weight Management** | 33,642 (61.0) | 3,091 (63.8) | <0.0001 | 8,168 (24.3) | 645 (20.9) | <0.0001 |
| BMI ≥30.0, n (%) | 24,467 (44.4) | 2,089 (43.1) | -- | 7,052 (28.8) | 553 (26.5) | -- |
| BMI 27.0-29.9 + Comorbidity, | 9,175 (16.6) | 1,002 (20.7) | -- | 1,116 (12.2) | 92 (9.2) | -- |
| Hypertension | 7,477 (13.6) | 786 (16.2) | -- | 620 (8.3) | 52 (6.6) | -- |
| Hyperlipidemia | 5,125 (9.3) | 789 (16.2) | -- | 267 (5.2) | 46 (5.8) | -- |
| Obstructive Sleep Apnea | 236 (0.4) | 193 (4.0) | -- | 1 (0.4) | 0 (0.0) | -- |
| Diabetes Mellitus | 2,811 (5.1) | 306 (6.3) | -- | 846 (30.1) | 71 (23.2) | -- |
| **Secondary Prevention of CVD<sup>c</sup>** | 7,126 (12.9) | 852 (17.6) | <0.0001 | 422 (5.9) | 31 (3.6) | 0.006 |
| **Any of the above** | 34,496 (62.6) | 3,199 (66.1) | <0.0001 | 7,988 (23.2) | 651 (20.4) | <0.001 |

<sup>a</sup>Glucagon-like peptide-1 receptor agonists
<sup>b</sup>Among those with an indication for a GLP-1 RA
<sup>c</sup>Adults aged 45 years or older with a BMI of ≥27 and a history of myocardial infarction or stroke.

## CONCLUSION

Relative to non-smokers, adults who smoke were more likely to have an indication for a GLP-1 RA, but less likely to receive a prescription. This finding was consistent across all indications for GLP-1 RAs. There is an opportunity to increase utilization of GLP-1 RAs for weight and disease management, and in doing so, increase access to GLP-1 RAs among adults who smoke. When considering the well-established negative effects of smoking on metabolic health and the risk of weight gain post-cessation, GLP-1 RAs hold promise for simultaneously addressing critical metabolic and smoking-related health challenges in adults who smoke with an existing indication. Given the emerging data on GLP-1 RAs for smoking cessation, consideration should be given to integrating behavioral support for smoking cessation with the GLP-1 RA treatment.

## Data Availability

Data are available upon reasonable request to the corresponding author.

## References

1. Bush T, Levine MD, Deprey M, et al. Prevalence of weight concerns and obesity among smokers calling a quitline. Journal of smoking cessation. 2009;4(2):74–78.

2. Chiolero A, Faeh D, Paccaud F, Cornus J. Consequences of smoking for body weight, body fat distribution, and insulin resistance. The American journal of clinical nutrition. 2008;87(4):801–809.

3. Mambo A, Yang Y, Mahulu E, Zihua Z. Investigating the interplay of smoking, cardiovascular risk factors, and overall cardiovascular disease risk: NHANES analysis 2011–2018. BMC Cardiovascular Disorders. 2024;24(1):193.

4. Kim Y. The effects of smoking, alcohol consumption, obesity, and physical inactivity on healthcare costs: a longitudinal cohort study. BMC Public Health. 2025;25(1):873.

5. Yammine L, Green CE, Kosten TR, et al. Exenatide adjunct to nicotine patch facilitates smoking cessation and may reduce post-cessation weight gain: a pilot randomized controlled trial. Nicotine and Tobacco Research. 2021;23(10):1682–1690.

